# Adverse childhood family environment is associated with altered cardiovascular regulation during exercise among young adults in the Coronary Artery Risk Development in Young Adults (CARDIA) Study

**DOI:** 10.64898/2026.04.24.26351696

**Authors:** Nathaniel D.M. Jenkins, Austin T. Robinson, Bjoern Hornikel, Kara M. Whitaker, David R. Jacobs, Kiarri Kershaw, Kelley Pettee Gabriel

## Abstract

The purposes of this study were to determine whether adverse childhood family environment (ACFE) exposure is associated with altered hemodynamic responses to graded exercise in young adulthood, and whether this association was modified by sex and race in a large, population-based cohort. We hypothesized that ACFE exposure would be associated with an exaggerated exercise pressor response in young adulthood, independent of resting BP. We further hypothesized that the association between ACFE and the hemodynamic response to exercise would be stronger in females than males, and in Black versus White participants.

**Methods:** Exercising blood pressure (BP) and heart rate (HR) responses were recorded during graded exercise testing and ACFE exposure was assessed among 3,417 young adults (mean age = 25 ± 4 y; 44% female; 46% Black). Linear mixed-effects models that included participant-specific random intercepts and random slopes were used to assess the relation between ACFE exposure and exercising systolic (SBP), diastolic, and mean BP, pulse pressure (PP), pulse pressure index (PPI), heart rate (HR), and rate pressure product (RPP). All models were adjusted for resting values of the hemodynamic outcome, as well as age, sex, race, study center, body mass index, current hypertension medication use, smoking status, and alcohol consumption.

**Results:** Graded exercise hemodynamic responses were analyzed in 3,346–3,417 participants in the final models, providing 15,372–17,481 observations. Higher ACFE exposure was associated with lower SBP (β = −0.304 mmHg/ACFE, *p* = 0.033), HR (β = −0.485 bpm/ACFE, *p*<0.001), and RPP (β = −83.404 bpm·mmHg/ACFE, *p*=0.002) at the lowest workload, but steeper workload-related increases in SBP (interaction β = 0.044 mmHg·MET⁻¹·ACFE⁻¹, *p*=0.029), HR (β = 0.061 bpm·MET⁻¹·ACFE⁻¹, *p*<0.001), RPP (β = 10.16 bpm·mmHg·MET⁻¹·ACFE⁻¹, *p*=0.025), and PP (β = 0.052 mmHg·MET⁻¹·ACFE⁻¹, *p*=0.038) and PPI (β = 0.000232 units·MET⁻¹·ACFE⁻¹, *p*=0.018). These findings were robust to additional adjustment for central adiposity, exercise capacity, and maximal heart rate and heart rate recovery.

**Conclusion:** Our findings add nuanced evidence revealing that early adversity is associated with a demand-dependent shift in cardiovascular regulation, with attenuated responses at low demand, but more dramatic increases in pulsatile and myocardial load responses during progressive physiological stress.

## INTRODUCTION

Adverse childhood family environments (ACFEs) are characterized by emotional distant or unsupportive interactions, aggression, abuse, and neglect. These environments expose children to significant psychosocial stressors during sensitive developmental windows, and are thought to produce both activation and recalibration of allostatic regulatory systems.^1^ Thus, exposure to ACFEs is consistently and independently related to higher risk for hypertension, cardiovascular disease (CVD), and premature mortality from any cause among middle-aged adults.^2–6^ Regarding physiological mechanisms for the increased cardiovascular risk, young adults exposed to ACFE demonstrate impaired endothelium-dependent vasodilatory function.^7,8^ Notably, these impairments precede overt signs or symptoms of CVD ^9^ and exaggerated age-related increases in resting blood pressure (BP), which appear to manifest beginning in the fourth decade of life.^10^ Together, these findings suggest that ACFE exposure is a latent CVD risk factor, which can be uncovered as subclinical impairments in cardiovascular regulation in response to physiological stressors.

Exaggerated exercise pressor responses can be observed prior to overt elevations in resting BP, may reflect intermittent BP responses to daily stressors that produce target organ damage,^11^ and thus provide additive prognostic information above baseline BP for future clinical hypertension,^12^ CVD, and all-cause mortality risk.^13–15^ Indeed, the *cardiovascular reactivity hypothesis* posits that exaggerated cardiovascular responsiveness to stress promotes maladaptive vascular and ventricular adaptations that may lead to an increased risk for CVD.^16,17^ However, recent evidence from the Dunedin Multidisciplinary Health and Development Study and the Midlife in the United States (MIDUS) Study suggest that early life adversity is associated with reduced cardiovascular reactivity to generalized mental stressors (Stroop task and mental arithmetic) in middle-aged adults, supporting the ‘*blunting hypothesis of cardiovascular reactivity*’.^18^ Prior studies exploring the impacts of early life adversity on cardiovascular stress reactivity have deployed different stressors, and it is not clear whether or how ACFE exposure influences the exercise BP response. In addition, while prior work has primarily focused on systolic and/or mean BP and heart rate, inclusion of pulse pressure (PP), proportional pulse pressure or pulse pressure index (PPI), and rate-pressure product (RPP) may provide complementary insights into early alterations in cardiovascular regulation during exercise by indexing pulsatile load and vascular compliance and myocardial oxygen demand.^19–22^

Adverse childhood experiences are more prevalent in females,^23^ and there is evidence that the female HPA axis may be more vulnerable to programming in response to early-life stress.^24^ Furthermore, individuals belonging to racial and ethnic minority populations are more likely to be exposed to ACFEs, which are also more likely to co-exist with additional community level adversity and historical trauma, such that the effects of ACFEs may be more potent in underrepresented ethnic or racial minority groups leading to increased prevalence of adverse cardiovascular health outcomes throughout life. However, as noted by the American Heart Association, race and ethnicity have rarely been examined as a potential modifier of the relationship between early life adversity and cardiovascular outcomes.^25^

The Coronary Artery Risk Development in Young Adults (CARDIA) study provides a unique opportunity to examine the association between childhood environment and the pressor response to a graded physiological stressor (i.e., graded exercise) in young adulthood. Given the developmental focus of the present study, we examined exercise hemodynamic responses at the Year 0 CARDIA examination when participants were young adults and largely free of overt CVD, allowing us to examine early regulatory phenotypes that may confer cardiovascular risk, rather than hemodynamic alterations secondary to established disease. Because participants in CARDIA were balanced for sex and race at study enrollment, there was also a unique opportunity to examine the influence of sex and race on this association.

Therefore, the purposes of this study were to determine whether ACFE exposure is associated with altered hemodynamic responses to graded exercise in young adulthood, and whether this association was modified by sex and race in a large, population-based, diverse cohort. We hypothesized that individuals who grew up in harsher, riskier early life environments would display an exaggerated exercise pressor response in young adulthood, independent of resting BP. We further hypothesized that the association between childhood environment and the exercise pressor response would be stronger in females than males, and in Black versus White participants.

## METHODS

### Participants

The CARDIA study is an ongoing, prospective longitudinal, multi-site cohort study of 5,115 self-identified Black and White adults between the ages of 18 and 30 years who were recruited to complete an initial in-person clinical exam in 1985-1986 in Birmingham, AL, Chicago, IL, Minneapolis, MN, or Oakland, CA. Follow-up includes semi-annual contacts and in person examinations approximately every 2-5 years since baseline. In this study, participants were included if they attended the year 15 (2000-2001) CARDIA examination and completed a questionnaire assessing ACFEs (n=3,655), and if they also had exercise BP and covariate data from the year 0 CARDIA examination, resulting in a total sample size of 3,417 participants for this analysis (**Table 1**). Written informed consent was obtained from all participants at each exam, and the study procedures were reviewed and approved by the IRB at each center.

**Table 1.**
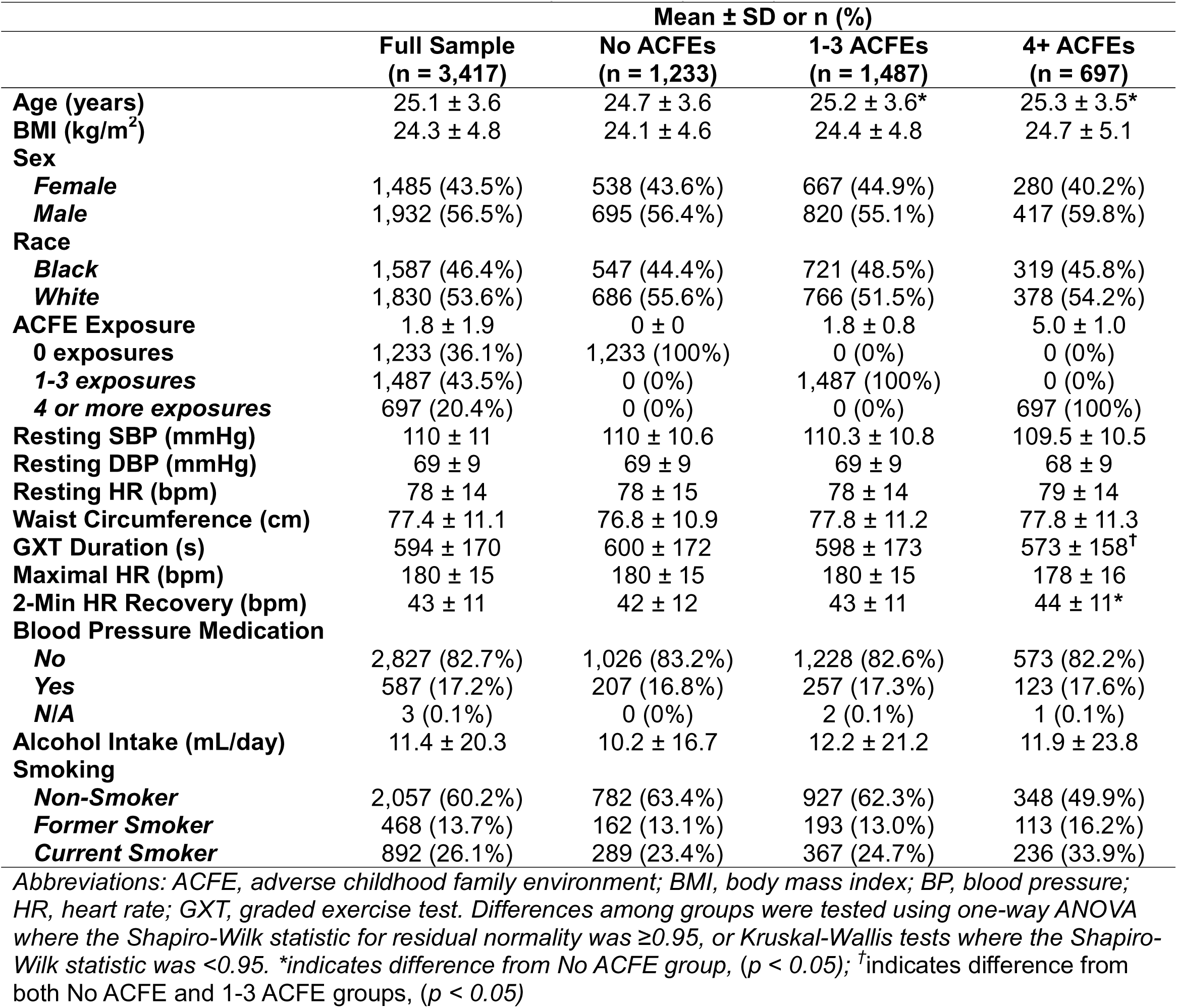
Participant characteristics for the full analytic sample (n=3,417).

### Assessment of ACFEs

ACFEs were retrospectively reported at the year 15 (2000-2001) CARDIA examination via in-person administration of a 7-item Risky Families questionnaire,^26^ which was adapted from the original ACFE questionnaire.^27^ Specifically, participants indicated the frequency at which they experienced each of the following 7 aspects of the family environment in childhood: parental love and support, verbal abuse, physical affection, physical abuse, presence of alcohol/drug abuser in the home, how well-organized and well-managed the household was, and parental/guardian knowledge of what participants were up to during childhood.^28^ We did not assume that each incremental increase on the Likert scale reflected a proportional exposure to childhood adversity because of the dissimilar degrees of severity across items. Instead, we utilized the scoring approach described by Pierce et al. ^28^ where each item was empirically dichotomized on the basis of the severity of psychosocial stress associated with each item. Accordingly, questions regarding physical, verbal, or emotional abuse, neglect, and exposure to individuals abusing drugs or alcohol were dichotomized as an exposure if participants reported any level of exposure. All other items were dichotomized at the mid-point of the 4-point Likert scale. Scores were then summated providing a range of possible exposure from 0 – 7 and this approach demonstrates strong internal consistency (Cronbach’s α = 0.73 – 0.76).^28^ The resulting distribution of ACFE exposure (none = 36.1%, 1-3 = 43.5%, high = 20.4%) is consistent with the distribution of adverse childhood experiences expected in the population.^29,30^

### Exercising Hemodynamic Responses

At the year 0 assessment, systolic blood pressure (SBP) and diastolic blood pressure (DBP) were recorded during a graded, maximal, symptom-limited exercise test on a treadmill by trained technicians using a random-zero sphygmomanometer during each stage of the exercise test. The exercise test was a Balke-type protocol, beginning at 3.0 mph and 2.0% grade (stage 1; 4.1 METs) and increasing progressively every two minutes by ∼1.8–1.9 METs, where the final possible stage was 5.6 mph and 25% grade (stage 9; 19.0 METs). Blood pressure was measured at 90s into each stage. While the rate of change in these blood pressures across workload during graded exercise tests have been shown to be predictive of future cardiovascular outcomes,^31–35^ the BP rate of change during the exercise test has not been explored or reported from CARDIA. Data were examined for non-physiologic values (i.e., SBP ≤70 mmHg; DBP ≤ 40 mmHg) and non-physiologic observations were removed from analysis. To then establish viability of the BP data, we first examined the association of BP and workload (stage) to assess the variability and degree to which the SBP, mean blood pressure 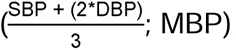, and pulse pressure increased monotonically and DBP decreased modestly with increases in workload. After confirming BP changes were as expected with increasing exercise workload (Figure 1), we defined the rates of change for SBP, MBP, DBP, HR, pulse pressure (PP), pulse pressure index 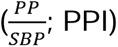, and rate pressure product (HR x SBP; RPP^20^) as the primary outcomes of interest.

**Figure 1.**
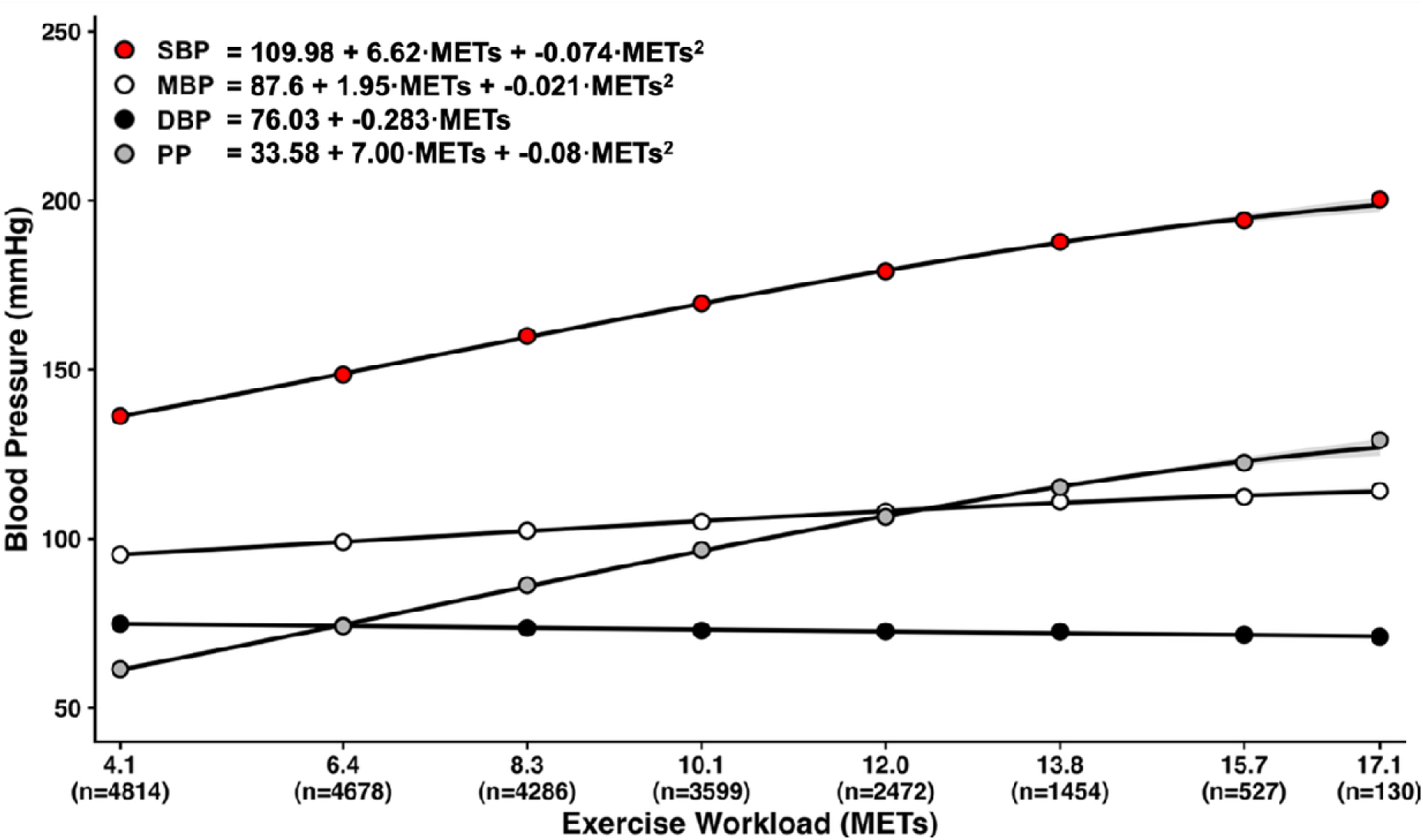
Blood pressure versus metabolic equivalents of task (METs) relationships in the full sample. Systolic blood pressure (R^2^ = 0.40, *p* < 0.001), mean blood pressure (R^2^ = 0.18, *p* < 0.001), and pulse pressure (R^2^ = 0.39, *p* < 0.001) increased monotonically with increasing workload. Diastolic blood pressure (R^2^ = 0.01, *p* < 0.001) decreased modestly with increasing workload. Sample size for each stage is shown below each MET value on the x-axis. *Abbreviations: DBP, diastolic blood pressure; MBP, mean blood pressure; METs, metabolic equivalents of task; PP, pulse pressure; SBP, systolic blood pressure*.

### Covariates

All covariates were assessed at the year 0 examination concurrent with completion of exercise testing. Height and weight were measured by trained technicians with participants wearing socks, shorts, a short-sleeved shirt or blouse, and no shoes. Height was measured to the nearest half centimeter, while weight was measured to the nearest 0.2 pounds using a calibrated balance scale. Body mass index (BMI) was derived from height and weight in kg/m^2^. Waist circumference was calculated as the average of two measurements (to the nearest 0.5 cm) performed at the level midway between the iliac crest and the lowest lateral portion of the rib cage using a cloth anthropometric tape. Cigarette smoking status was assessed via a combination of self-reported and interviewer-administered follow-up questions to classify participants as never, former, or current smokers. Antihypertensive medication use was assessed via an interviewer-administered medication inventory in which participants were asked to prepare and bring a list of current medications to the examination. Alcohol consumption was assessed via an interviewer-administered questionnaire that captured both the frequency and quantity of intake over the past year, from which an average daily ethanol intake (mL/day) was derived.

### Statistical Analyses

ACFE exposure was quantified as described above and modeled as a continuous predictor. Exercising hemodynamic responses (SBP, MBP, DBP, PP, PPI, RPP) were analyzed using linear mixed-effects models to account for repeated measurements obtained during graded treadmill exercise testing. The hemodynamic variables measured at each exercise stage served as the dependent variable with exercise workload (i.e., METs) as the within participant, intensity-varying predictor. Exercise stage workload was treated as a continuous variable and centered at the workload during the first exercise stage (4.1 METs) to facilitate physiological interpretation of model intercepts and main effects. The base models included fixed effects for METs, ACFE, and their interaction (METs×ACFE), and adjusted for resting values of the hemodynamic outcome. Models also included age, sex, race, study center, body mass index (BMI), current hypertension medication use, smoking status, and alcohol consumption as covariates, which were all selected a priori based on their physiological relevance. Sex, race, and study center were included as categorical factors. ACFE and all continuous covariates were mean-centered prior to analysis to aid interpretation of regression coefficients.

To account for within-participant correlation across exercise stages and inter-individual variability in hemodynamic reactivity, models included participant-specific random intercepts and random slopes as follows (index i is person and j is stage within person):

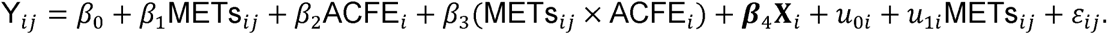

Random effects were assumed to follow a multivariate normal distribution and normal distribution and constant variance were assumed for residual errors. Models were estimated using restricted maximum likelihood (REML). Degrees of freedom for fixed-effect tests were calculated using the Satterthwaite approximation.

To evaluate whether adversity effects varied non-linearly across the exposure distribution, models incorporating natural cubic spline terms for ACFE were also compared with linear models using likelihood-ratio tests estimated via maximum likelihood. The likelihood-ratio test was not statistically significant for any outcome (all p > 0.05), and thus linear terms were used for ACFE. Similarly, linearity of exercise workload (METs) effects was evaluated using natural cubic spline models. While spline models generally improved statistical fits, visual inspection of the predicted values demonstrated near-linear associations across the observed workload range, with minimal notable deviation from linearity. Accordingly, linear parameterizations of workload were retained for parsimony and interpretability.

To test our hypotheses that ACFE-related differences in exercise hemodynamic responses would differ by sex and race, the base model was extended to include higher-order, three-way interaction terms (METs×ACFE×sex and METs×ACFE×race). Nested models were compared using likelihood-ratio tests estimated by maximum likelihood. The three-way interaction terms including sex and race were not significant. However, both sex and race significantly modified the hemodynamic-workload relationships and inspection of fixed-effect estimates indicated that improvements in model fits were primarily attributable to the METs×sex and METs×race interaction terms. Accordingly, we decided to retain METs×sex and METs×race interaction terms but excluded the 3-way terms for all final primary models, which were estimated using REML with Satterthwaite degrees-of-freedom approximation.

To aid interpretation of interaction effects, model-based predictions of the dependent variables across exercise intensity were generated for representative adversity levels corresponding to the 10th, 50th, and 90th percentiles of the centered adversity distribution and 95% confidence intervals were derived from the fixed-effects variance–covariance matrix using the delta method. The workload at which predicted hemodynamic responses converged across adversity levels was calculated from model coefficients and uncertainty (95% CI) was estimated using a parametric bootstrap with repeated sampling (1,000 replications) of fixed-effect coefficients from their multivariate normal distribution using the fixed-effects variance–covariance matrix. Marginal effects of ACFE on hemodynamic outcomes across exercise workloads were computed to assess whether adversity-related differences in hemodynamic outcomes became more pronounced at higher exercise intensities.

A sensitivity analysis was conducted by extending the final primary models to additionally adjust for waist circumference, exercise test duration, maximal heart rate, and two-minute heart-rate recovery.^36^ The purpose of these analyses was to evaluate whether ACFE-related differences in exercise hemodynamic responses were independent of central adiposity, exercise capacity, chronotropic competence, and autonomic recovery/fitness. These models were estimated using complete-case data and retained the same random-effects structure and covariate adjustment set as the primary models.

An alpha of 0.05 was considered statistically significant and all analyses were conducted in R (R Foundation for Statistical Computing, Vienna, Austria) using the lme4 and lmerTest packages. Base models were coded by the lead author, and code refinement and visualization formatting were assisted by ChatGPT (v5.2; OpenAI), and all analyses were verified by the authors. Data are reported as mean±SD, mean [95% CI LB, 95% CI UB], and β±SE unless otherwise specified.

## RESULTS

Graded exercise hemodynamic responses were analyzed in 3,346–3,417 participants in the final analytic models, providing between 15,372 and 17,481 exercise-stage observations dependent on the outcome. Thus, participants provided an average of 4.6±1.6 – 5.3±1.5 exercise stage observations for each outcome.

**Table 2** provides β coefficients ± SE for model terms of interest for each hemodynamic outcome. As expected, SBP increased progressively, MBP increased modestly, and DBP decreased modestly with increasing exercise workload. In addition, PP, PPI, HR, and RPP all increased robustly with increasing exercise workload (all p < 0.001).

**Table 2.** Results for the primary two-way mixed effects restricted maximum likelihood models. β coefficients ± SE are shown for model terms of interest. Each column represents a single model with the indicated hemodynamic outcome as the dependent variable. In addition to covariates shown (sex and race), models were adjusted for age, resting hemodynamic values, BP medication use, smoking, alcohol, BMI, and study center (full results shown in supplementary Table 1). Because METs were centered at the lowest exercise stage (4.1 METs), the ACFE main effect reflects the association between ACFE exposure and the hemodynamic outcome at the lowest workload, and the METs×ACFE interaction term reflects the degree to which the ACFE effect changes with each additional MET of exercise intensity. ^#^, p < 0.1; *, *p* < 0.05; ** *p* < 0.01; \*\*\**p* < 0.001.

|  | SBP | DBP | MBP | PP | PPI | HR | RPP |
| --- | --- | --- | --- | --- | --- | --- | --- |
| **Intercept** | 146.6 ± 0.7*** | 75.9 ± 0.41*** | 99.2 ± 0.39*** | 72.3 ± 0.77*** | 0.49 ± 0.003*** | 102.9 ± 0.59*** | 15276.2 ± 136.4*** |
| **METs** | 6.00 ± 0.07*** | -0.73 ± 0.05*** | 1.51 ± 0.04*** | 6.66 ± 0.09*** | 0.02 ± 0.0003*** | 8.57 ± 0.05*** | 2321.7 ± 16.2*** |
| **ACFE** | -0.30 ± 0.14* | -0.14 ± 0.08<sup>#</sup> | -0.22 ± 0.08** | -0.17 ± 0.15 | -0.001 ± 0.0007 | -0.49 ± 0.12*** | -83.4 ± 26.5** |
| **SEX** | -5.97 ± 0.61*** | -2.60 ± 0.33*** | -4.25 ± 0.33*** | -5.55 ± 0.63*** | -0.02 ± 0.003*** | 14.97 ± 0.49*** | 1553.0 ± 106.2*** |
| **RACE** | 0.72 ± 0.58 | -1.38 ± 0.32*** | -0.91 ± 0.32** | 1.54 ± 0.62** | 0.01 ± 0.003*** | -2.57 ± 0.48*** | -469.0 ± 108.0*** |
| **METs×ACFE** | 0.04 ± 0.02* | -0.001 ± 0.01 | 0.02 ± 0.01 | 0.05 ± 0.02* | 0.0002 ± 0.0001* | 0.06 ± 0.01*** | 10.2 ± 4.5* |
| **METs×Sex** | -0.35 ± 0.08*** | 0.34 ± 0.05*** | 0.14 ± 0.05** | -0.61 ± 0.09*** | 0.0006 ± 0.0004<sup>#</sup> | 0.44 ± 0.06*** | -126.6 ± 17.5*** |
| **METs×Race** | -0.86 ± 0.08*** | 0.21 ± 0.05*** | -0.14 ± 0.05** | -1.02 ± 1.0*** | -0.003 ± 0.0004*** | -0.20 ± 0.06*** | -168.3 ± 17.7*** |

Abbreviations: ACFE, adverse childhood family environment; BMI, body mass index; BP Medication, blood pressure medication use; METs, metabolic equivalents of task

Higher (less favorable) ACFE exposure was associated with lower SBP at the lowest workload (*p* = 0.033), and the METs×ACFE interaction was significant (*p* = 0.029), indicating steeper workload-provoked SBP increases among individuals with higher ACFE (**Figure 2**). ACFE exposure was also associated with lower DBP (*p* = 0.077) and MBP (*p* = 0.005) at the lowest workload, whereas the METs×ACFE interaction was not significant for either DBP (*p* = 0.94) or MBP (*p* = 0.15). ACFE exposure was not associated with PP (*p* = 0.27) or PPI (*p* = 0.88) at the lowest workload, but the METs×ACFE interaction was significant for both PP (*p* = 0.038) and PPI (*p* = 0.018) (**Figure 3**). Finally, ACFE exposure was associated with lower HR (*p* < 0.001) and RPP (*p* = 0.002) at the lowest workload, and the METs×ACFE interaction was significant for both HR (*p* < 0.001) and RPP (*p* = 0.025) (**Figure 4**).

**Figure 2.**
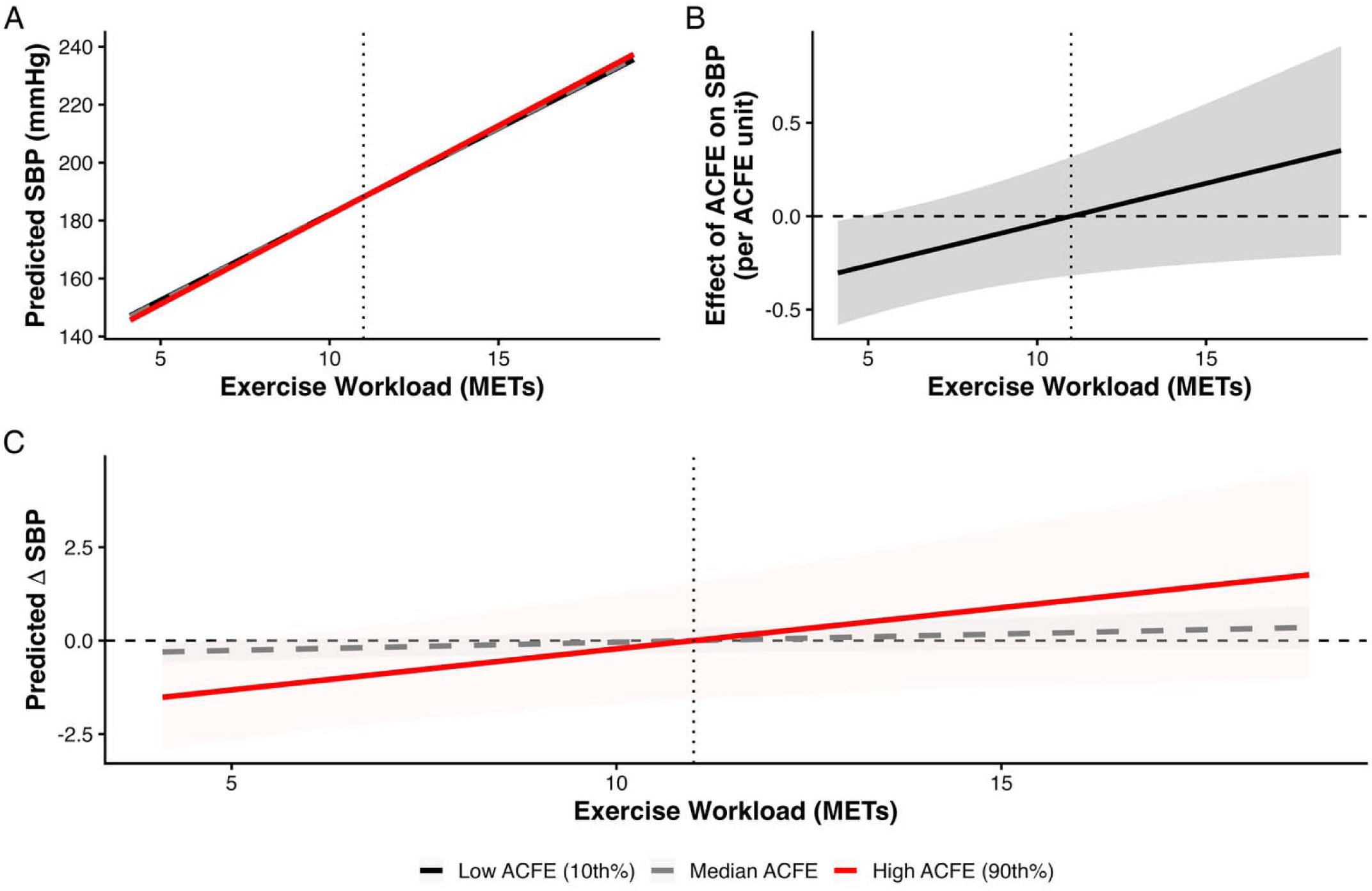
Systolic blood pressure responses to graded exercise intensity by adverse childhood family environment (ACFE) exposure. (**A**) Model-predicted systolic blood pressure (SBP) across exercise workload (METs) for low (10th percentile; 0 ACFEs), moderate (50th percentile; 1 ACFE), and high (90th percentile; 5 ACFEs) ACFE from linear mixed-effects models using participant-specific random intercepts and workload slopes. Note that Panel A illustrates the overall SBP trajectory across the exercise workload range; because absolute SBP increases by approximately 100 mmHg from rest to peak exercise, between-group differences at any given workload are difficult to discern visually at this scale and are better appreciated in Panels B and C. The vertical, dashed dotted line represents the estimated workload at which predicted SBP values for high versus low ACFE are equal (11.0 METs). (**B**) The estimated marginal association of ACFE with SBP (mmHg per ACFE unit) across exercise workload derived from the METs×ACFE interaction term. (**C**) The model-predicted SBP difference relative to low ACFE (ΔSBP; median – low ACFE (dashed grey line) and high – low ACFE (solid red line)) across workload computed from linear contrasts of model predictions. The horizontal dashed line indicates no difference from the low ACFE group. All models were adjusted for age, sex, race, study center, resting SBP, body mass index, hypertensive medication use, smoking, and alcohol use. Shaded bands represent 95% confidence intervals for all panels. *Note that in panel A,

**Figure 3.**
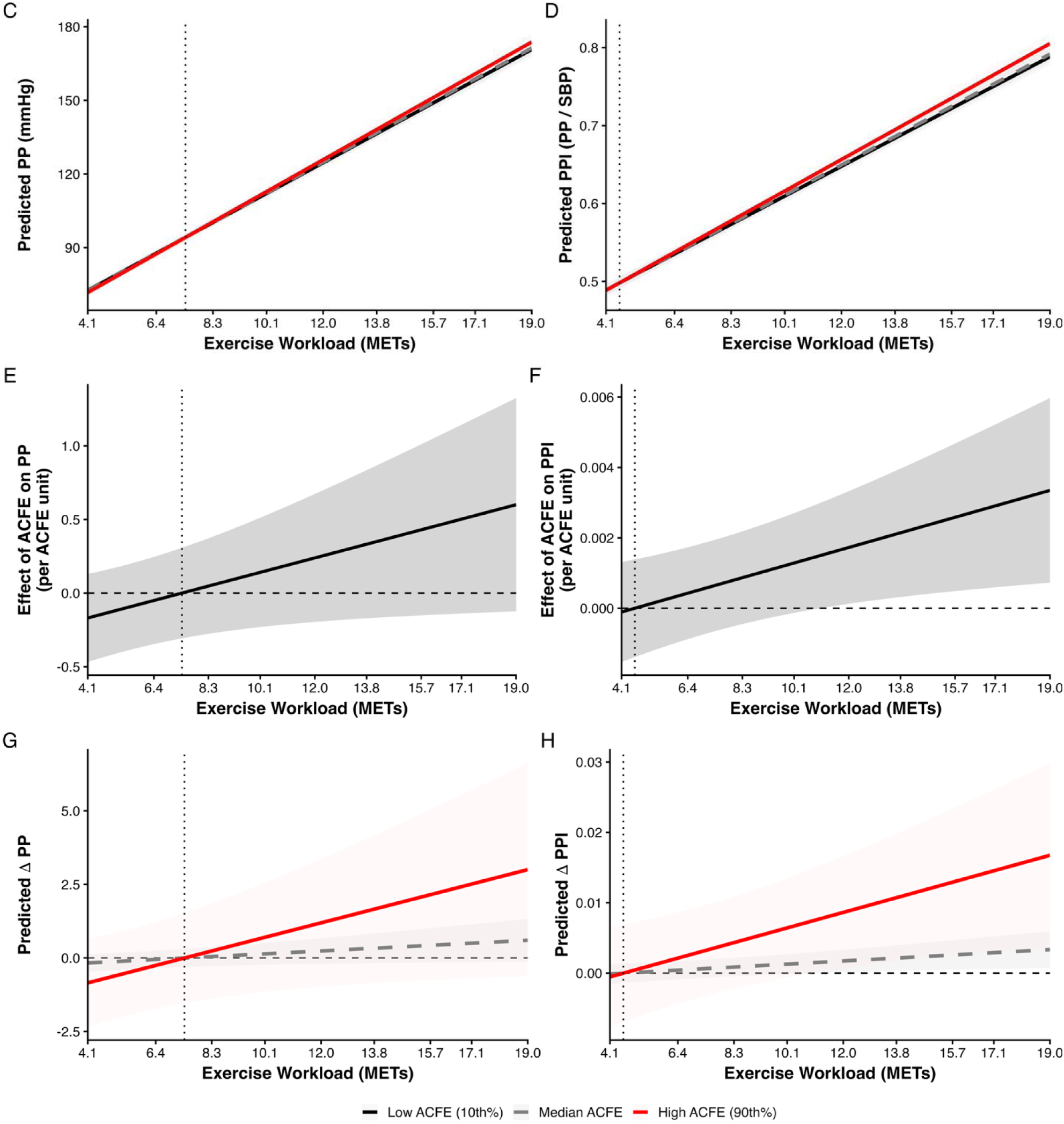
Pulsatile pressure (PP) and pulsatile pressure index (PPI) response to graded exercise intensity by adverse childhood family environment (ACFE) exposure. (**A** and **B**) Model-predicted PP and PPI across exercise workload (METs) for low (10th percentile; 0 ACFEs), moderate (50th percentile; 1 ACFE), and high (90th percentile; 5 ACFEs) ACFE from linear mixed-effects models using participant-specific random intercepts and workload slopes. The vertical, dashed dotted line represents the estimated workload at which predicted PP and PPI values for high versus low ACFE are equal (7.4 and 4.6 METs, respectively). (**C** and **D**) The estimated marginal association of ACFE with PP and PPI (mmHg per ACFE unit) across exercise workload derived from the METs×ACFE interaction term. (**E** and **F**) The model-predicted PP and PPI difference relative to low ACFE (ΔPP/PPI; median – low ACFE (dashed grey line) and high – low ACFE (solid red line)) across workload computed from linear contrasts of model predictions, where the horizontal dashed line indicates no difference from the low ACFE group. All models were adjusted for age, sex, race, study center, resting SBP, body mass index, hypertensive medication use, smoking, and alcohol use. Shaded bands represent 95% confidence intervals for all panels.

**Figure 4.**
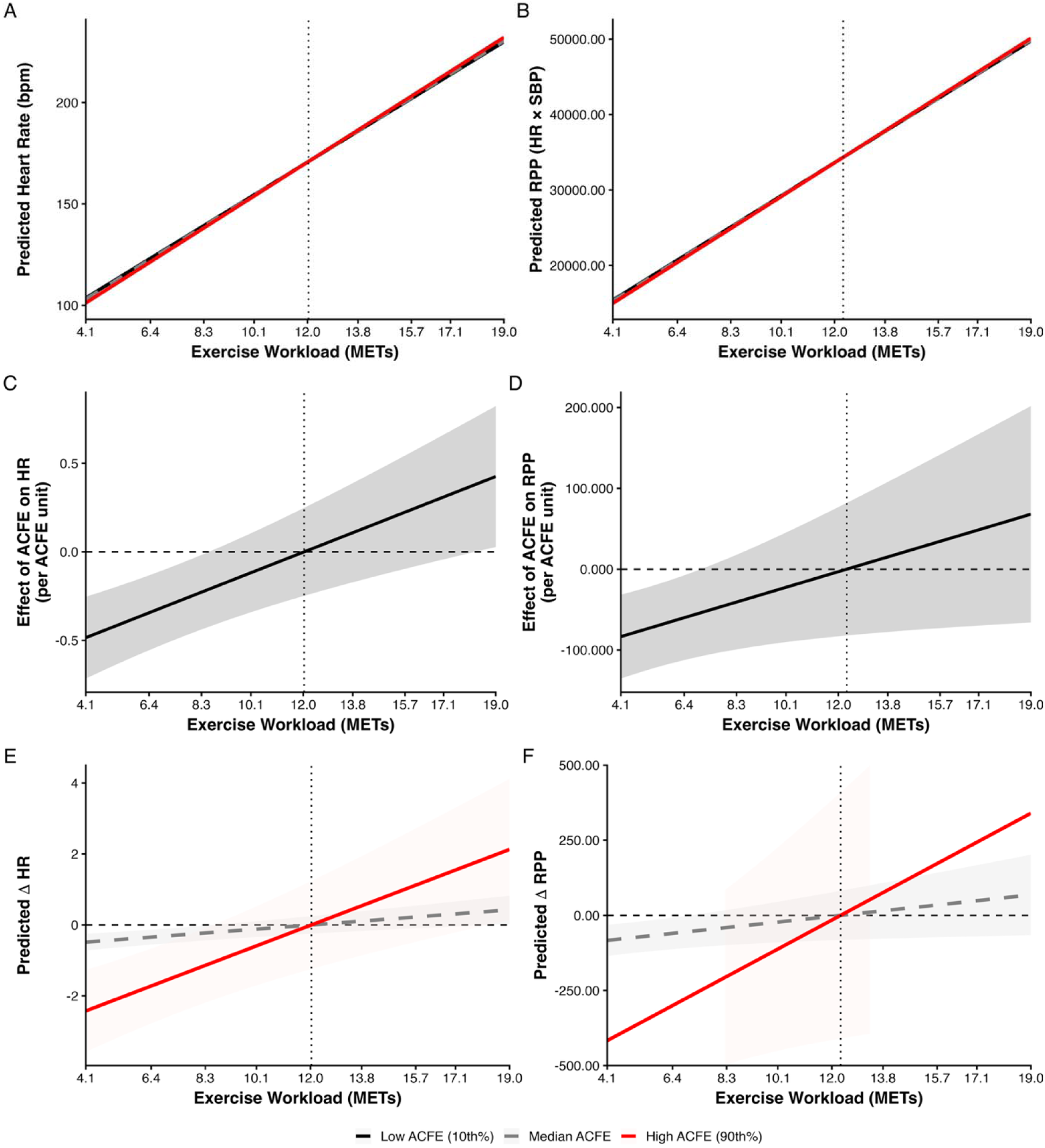
Heart rate (HR) and rate pressure product (RPP) response to graded exercise intensity by adverse childhood family environment (ACFE) exposure. (**A** and **B**) Model-predicted HR and RPP across exercise workload (METs) for low (10th percentile; 0 ACFEs), moderate (50th percentile; 1 ACFE), and high (90th percentile; 5 ACFEs) ACFE from linear mixed-effects models using participant-specific random intercepts and workload slopes. The vertical, dashed dotted line represents the estimated workload at which predicted HR and RPP values for high versus low ACFE are equal (12.0 and 12.3 METs, respectively). (**C** and **D**) The estimated marginal association of ACFE with HR and RPP (mmHg per ACFE unit) across exercise workload derived from the METs×ACFE interaction term. (**E** and **F**) The model-predicted HR and RPP difference relative to low ACFE (ΔPP/PPI; median – low ACFE (dashed grey line) and high – low ACFE (solid red line)) across workload computed from linear contrasts of model predictions, where the horizontal dashed line indicates no difference from the low ACFE group. All models were adjusted for age, sex, race, study center, resting SBP, body mass index, hypertensive medication use, smoking, and alcohol use. Shaded bands represent 95% confidence intervals for all panels.

Model-based contrasts relative to low ACFE exposure demonstrated that predicted differences between ACFE levels crossed zero for SBP at 11.0 [5.7, 17.8] METs, for HR at 12.0 [8.7, 17.0] METs, and for RPP at 12.3 [7.1, 18.2] METs, and became progressively greater at higher workloads. While predicted crossover points were also able to be identified for PP (7.4 [4.5, 15.9] METs), PPI (4.6 [4.2, 12.2] METs), and MBP (17.1 [7.4, 18.7]), these were less meaningful because there were either no significant ACFE-related differences observed at low-intensities (e.g., PP and PPI) or no significant ACFE-related differences in the rate of change observed across intensity (e.g., MBP).

In sum, these findings suggest that individuals with higher ACFE exhibited lower predicted SBP, HR, and RPP at lower exercise workloads compared to those with lower ACFE, whereas workload-related SBP, HR, and RPP increases were steeper among those with higher ACFE, resulting in convergence of predicted values at moderate-to-vigorous intensity workloads and divergence at vigorous intensity workloads.

### Sex- and Race-Related Moderation Effects

Both sex and race significantly modified workload responses across multiple outcomes. For SBP and RPP, the workload-related increases were smaller in females than males (METs×sex; SBP: *p* = 6.16×10⁻□; RPP: *p* < 0.001). For HR, the workload-related increase was greater in females (METs×sex; *p* < 0.001). Race similarly modified workload responses for SBP (METs×race; *p* < 0.001) and RPP (*p* < 0.001), indicating that workload-related increases were lower in White than in Black participants. These findings indicate consistent sex- and race-related differences in workload-dependent cardiovascular responses during graded exercise, independent of ACFE. Notably, sex did not modify the METs×ACFE relation for any of the hemodynamic outcomes, and race only modified the METs×ACFE relation for HR (F_1, 2988.8_ = 5.05, *p* = 0.025). The ACFE-related augmentation of the HR response to increasing workload was stronger among White than Black participants (β = 0.07±0.03 bpm per MET per ACFE unit; **Supplementary Figure 1**).

### Sensitivity Analyses for Fitness, Central Adiposity, and Heart Rate Recovery

Sensitivity analyses were conducted to determine whether associations between ACFE and exercise hemodynamic responses were explained by differences in adiposity, cardiorespiratory fitness, or autonomic recovery. Models were additionally adjusted for waist circumference, graded exercise test duration, maximal heart rate, and two minute heart-rate recovery. **Supplementary Table 2** provides β coefficients ± SE for model terms of interest for each hemodynamic outcome for the sensitivity analyses. Across outcomes, inclusion of these variables did not materially alter the primary findings. For SBP, the METs×ACFE interaction remained significant (*p* = 0.034), although the association between ACFE and SBP at the lowest workload was attenuated and no longer statistically significant (*p* = 0.056). Similarly, workload-dependent ACFE associations persisted for HR (*p* < 0.001), RPP (*p* = 0.030), PP (*p* = 0.039), and PPI (β = 0.0002 MET⁻¹·ACFE⁻¹, *p* = 0.020). The METs×ACFE interactions for DBP (*p* = 0.919) and MBP (*p* = 0.169) remained non-significant after adjustment. Together, these findings indicate that the differences in exercise hemodynamic responses associated with ACFE were not explained by central adiposity, exercise capacity, maximal chronotropic response, or early post-exercise autonomic recovery.

## DISCUSSION

To our knowledge, this is the first study to examine associations between adverse childhood family environment (ACFE) and cardiovascular responses to graded exercise in a large population-based cohort. Three notable findings provide important insight into the effects of early life adversity on cardiovascular regulation in young adulthood and provide important context for interpreting prior studies reporting both attenuated ^18,37–39^ and exaggerated ^40–42^ cardiovascular responses following early life stress. First, ACFE was associated with attenuated blood pressure and heart rate responses at low exercise intensities (**Table 2**, **Figure 2C**, **Figure 4E/F**). Second, ACFE was associated with greater workload-dependent increases in pulsatile pressure and myocardial demand responses, as reflected by greater increases in SBP, PP, PPI, and RPP with increasing exercise intensity (**Figures 2-4**). The identification of crossover workloads for SBP and related measures further indicates that ACFE-related differences are not static, but rather evolve across increasing cardiovascular demand. Third, there were no notable moderating effects of either sex or race on the ACFE versus hemodynamic relations observed in this study, except that the association of ACFE on workload-related increases in heart rate were stronger in White than Black adults (**Supplementary Figure 1**). It is also notable that the observed patterns were not materially changed by additional adjustment for indicators of central adiposity, exercise capacity, chronotropic competence, or heart rate recovery. Together, our findings suggest that early adversity produces a phenotype that reduces cardiovascular activation during lower intensity activity, but is adversely associated with cardiovascular reserve capacity, reducing buffering capacity when cardiac output and pulsatile load increase. Thus, rather than reflecting uniformly lower or higher cardiovascular reactivity, the present findings suggest that early adversity may be characterized by a shift in the regulation of cardiovascular responses across the physiological demand spectrum, with relatively attenuated responses at low demand but potentially exaggerated pulsatile and myocardial load responses during more intense physiological stress.

One of the primary findings of the present study was that greater ACFE exposure was associated with reduced cardiovascular reactivity at low-to-moderate exercise workloads. This finding is in opposition to the cardiovascular reactivity hypothesis, which suggests that increased cardiovascular stress reactivity is a mediating pathway by which ACFEs are biologically embedded to pathophysiological outcomes, such as vascular dysfunction and ventricular remodeling, which promote poor cardiovascular health outcomes.^18,43–46^ In agreement with our findings, however, Bourassa *et al.* ^18^ reported that greater early life adversity was associated with lower cardiovascular reactivity to mental stress in independent longitudinal cohort studies of young (Dunedin Multidisciplinary Health and Development Study) and middle-aged (MIDUS Study) adults. Notably, lower cardiovascular reactivity was linked to accelerated biological aging or premature mortality in these studies. Similarly, McLaughlin and colleagues ^37^ reported that childhood maltreatment was associated with reduced cardiac output and elevated total peripheral resistance reactivity to the Trier Social Stress test. Given the nature of mental stressors and the potential for individual differences in factors such as task engagement or conscientiousness to alter stress responses, Bourassa and colleagues ^18^ highlighted the need for future studies to deploy stressful tasks that are robust to these individual factors. Thus, our study represents an important extension of this work, allowing us to assess cardiovascular stress reactivity to a generalized, intensity-varying physiological stressor (i.e., graded exercise). Our data support that increasing ACFE exposure is associated with lower blood pressure and heart rate reactivity at low-to-moderate workloads. Together, these findings are consistent with and lend support for the blunting hypothesis of cardiovascular reactivity and suggest that this blunting is, at least in part, physiological and not necessarily dependent on individual level psychological factors.

In the present study, ACFE was associated with steeper workload-dependent increases in SBP, PP, PPI, and RPP. The cardiovascular response to dynamic exercise reflects integrated actions of central command, the baroreflexes, and the exercise pressor reflex, alongside peripheral vascular regulatory mechanisms that modulate resistance, compliance, and pulse wave reflection as cardiac output increases. Exercise-related increases in cardiac output and blood flow elevate endothelial shear stress, promoting vasodilation and functional increases in arterial compliance. These physiological vascular responses promote reductions in wave reflection and serve to buffer the transmission of pulsatile energy as stroke volume rises, which constrains increases in systolic and pulsatile pressures.^47^ Consequently, impaired peripheral vasodilation also promotes greater pulse wave reflection and augmentation pressure during exercise, increasing total systolic pressure and myocardial workloads.^47,48^ Notably, recent studies have reported endothelial dysfunction among otherwise healthy young adults exposed to early life adversity, even in the absence of traditional cardiometabolic risk factors.^7,49–51^ Exaggerated pulsatile pressure responses may also reflect altered ventricular–vascular coupling, which can be disrupted by insufficient vascular buffering. For example, reductions in arterial compliance increase effective arterial elastance, or ventricular afterload.^52^ If changes in arterial elastance are not matched by proportional augmentation of ventricular end-systolic elastance during stress, higher systolic pressures must be generated to overcome the increase in arterial load resulting in amplified pulsatile pressure and myocardial workload increases with increasing exercise intensity. Notably, these effects could result from either disproportionate increases in arterial load and/or limited ventricular reserve.^53,54^ Thus, systemic endothelial dysfunction and altered ventricular–vascular coupling together provide a unifying physiological explanation for the observed workload-dependent increases in SBP, PP, PPI, and RPP among individuals with greater ACFE exposure in the present study. This explanation is further supported by the fact that our findings were robust to additional adjustment for waist circumference, exercise capacity, maximal heart rate, and heart rate recovery, suggesting that differences in adiposity, fitness, and potentially autonomic function (i.e., parasympathetic reactivation/sympathetic withdrawal) do not explain the association.

Importantly, this dynamic pattern also aligns with emerging models of endothelial and ventricular–vascular dysfunction in which early, latent abnormalities are revealed when cardiovascular regulatory systems are challenged.^7,54^

Taken together, our findings suggest that early life adversity is associated with a demand-dependent shift in cardiovascular regulation. The blunted responses observed at lower workloads are consistent with a growing body of literature demonstrating attenuated physiological responses to mental stressors following early life adversity,^55,56^ reflecting altered central stress-response regulation (i.e., blunted HPA- and SAMs-axes activation) that may result in reduced neurocardiovascular activation, particularly during low-demand conditions. However, our finding of exaggerated demand-dependent increases in systolic and pulse pressures, alongside heart rate and myocardial oxygen demand, may reflect diminished endothelial buffering and altered ventricular–vascular interaction, which would be expected to be increasingly consequential as cardiac output rises. Together, these findings suggest multi-level recalibration of neurovascular regulatory systems that reduce regulatory flexibility and limit the dynamic range over which cardiovascular responses can be adjusted to meet changing demands. Future studies utilizing higher fidelity measurements will be necessary to test this hypothesis. However, this premise is consistent with developmental programming and allostatic load models whereby early life environmental stress is thought to recalibrate stress-responsive systems and/or reduce the energetic capacity for growth and maintenance of physiological systems,^57–59^ biasing neurocardiovascular activation toward conservation during low-demand at the expense of reducing the capacity to accommodate escalating hemodynamic requirements. Importantly, even subtle workload-dependent increases in systolic and pulsatile pressures during young adulthood may have meaningful long-term consequences. Indeed, repeated exposure to exaggerated pulsatile pressures and myocardial workloads during routine stress could accelerate endothelial injury, promote arterial stiffening, and reduce myocardial efficiency over time. Thus, a compressed regulatory functional range may not manifest overtly as resting dysfunction but could predispose individuals to premature development of CVD. Finally, although not directly assessed, cellular bioenergetic dysfunction could contribute to both altered central stress-responses and impaired vascular and cardiac function, representing a potential unifying mechanistic, molecular link that warrants systematic investigation.^7,58,60^ In summary, the modest differences observed in the present study are likely to represent early pathophysiological signatures that contribute to lifetime cardiovascular risk.

Major strengths of the present study include the large, population-based sample and comprehensive examination of hemodynamic responses across the broad range of the physiological, cardiovascular demand spectrum, representing a first of its kind study. Another strength includes the use of hierarchial mixed-effects modeling that accounted for within-person clustering, inter-individual variability in reactivity, physiologically-relevant covariate adjustment. The additional sensitivity and model-specification analyses further support the stability of our findings. Several limitations should also be considered when interpreting these findings. Assessments of ACFE were conducted at year 15 and introduce the potential for recall bias or differential reporting secondary to psychosocial factors in adulthood. While ACFE itself was temporally antecedent to exercise testing at year 0, the retrospective self-report may not perfectly capture early life environment(s). It is notable that ACFE exposure has been associated with CVD incidence over 30 years of follow-up in the CARDIA study.^28^ Blood pressures were also obtained by manual auscultation due to technological standards at the time of assessment, which limits mechanistic inference regarding wave reflection, arterial elastance, or central pressures. Finally, despite conducting hypothesis driven covariate adjustment, hierarchical modeling, and additional sensitivity analysis, we cannot exclude the possibility of residual confounding.

In conclusion, our findings add nuanced evidence revealing that early adversity is associated with a demand-dependent shift in cardiovascular regulation, whereby responses at low demand are generally attenuated, but pulsatile and myocardial load responses increase more dramatically during progressive physiological stress. These findings are consistent with altered central-stress regulatory responsiveness and reduced vascular buffering capacity that together may impair the capacity to accommodate increasing hemodynamic demands during exercise. Moreover, the fact that these patterns were largely consistent across race and sex suggest that this demand-dependent cardiovascular phenotype is broadly observed following early life adversity and is not strongly modified by race or sex, although the stronger association between ACFE exposure and workload-dependent increases in heart rate among White than Black adults may deserve further investigation. Therefore, these data provide an important framework for understanding how early life adversity may be associated with (mal)adaptive cardiovascular regulatory dynamics during stress and may inform efforts aimed at early risk detection and pursuit of intervenable targets to reduce this risk.

## Data Availability

Data are available upon reasonable request in accordance with policy and procedures described in the CARDIA Study Data and Materials Distribution Agreement.

## ACKNOWLEDGEMENTS

The Coronary Artery Risk Development in Young Adults Study (CARDIA) is conducted and supported by the National Heart, Lung, and Blood Institute (NHLBI) in collaboration with the University of Alabama at Birmingham (75N92023D00002 & 75N92023D00005), Northwestern University (75N92023D00004), University of Minnesota (75N92023D00006), and Kaiser Foundation Research Institute (75N92023D00003). Funding was additionally supported by the CARDIA Fitness Study (R01HL078972). This manuscript has been reviewed by CARDIA for scientific content.

## Abbreviations and Acronyms

ACFE: Adverse Childhood Family Environment
ACE: Adverse Childhood Experience
BMI: Body Mass Index
BP: Blood Pressure
CARDIA: Coronary Artery Risk Development in Young Adults
CI: Confidence Interval
CVD: Cardiovascular Disease
DBP: Diastolic Blood Pressure
GXT: Graded Exercise Test
HPA: Hypothalamic-Pituitary-Adrenal (axis)
HR: Heart Rate
MBP: Mean Blood Pressure
METs: Metabolic Equivalents (of Task)
MIDUS: Midlife in the United States Study
PP: Pulse Pressure
PPP: Proportional Pulse Pressure
REML: Restricted Maximum Likelihood
RPP: Rate Pressure Product
SAM: Sympathetic-Adreno-Medullary (axis)
SBP: Systolic Blood Pressure
SE: Standard Error

---

**Supplementary Table 1.** Full results for the primary two-way mixed effects restricted maximum likelihood models. β coefficients ± SE are shown for each model term. In addition to co-variates shown, models were adjusted for study center (not shown). Because METs were centered at the lowest exercise stage (4.1 METs), the ACFE main effect reflects the association between ACFE exposure and the hemodynamic outcome at the lowest workload, and the METs×ACFE interaction term reflects the degree to which the ACFE effect changes with each additional MET of exercise intensity. ^#^, p < 0.1; *, *p* < 0.05; ** *p* < 0.01; \*\*\**p* < 0.001.

|  | SBP | DBP | MBP | PP | PPI | HR | RPP |
| --- | --- | --- | --- | --- | --- | --- | --- |
| **Intercept** | 146.6 ± 0.7*** | 75.9 ± 0.41*** | 99.2 ± 0.39*** | 72.3 ± 0.77*** | 0.49 ± 0.003*** | 102.9 ± 0.59*** | 15276.2 ± 136.4*** |
| **METs** | 6.00 ± 0.07*** | -0.73 ± 0.05*** | 1.51 ± 0.04*** | 6.66 ± 0.09*** | 0.02 ± 0.0003*** | 8.57 ± 0.05*** | 2321.7 ± 16.2*** |
| **ACFE** | -0.30 ± 0.14* | -0.14 ± 0.08<sup>#</sup> | -0.22 ± 0.08** | -0.17 ± 0.15 | -0.001 ± 0.0007 | -0.49 ± 0.12*** | -83.4 ± 26.5** |
| **SEX** | -5.97 ± 0.61*** | -2.60 ± 0.33*** | -4.25 ± 0.33*** | -5.55 ± 0.63*** | -0.02 ± 0.003*** | 14.97 ± 0.49*** | 1553.0 ± 106.2*** |
| **RACE** | 0.72 ± 0.58 | -1.38 ± 0.32*** | -0.91 ± 0.32** | 1.54 ± 0.62** | 0.01 ± 0.003*** | -2.57 ± 0.48*** | -469.0 ± 108.0*** |
| **Age** | -0.14 ± 0.07<sup>#</sup> | 0.19 ± 0.04*** | 0.08 ± 0.04* | -0.22 ± 0.08** | -0.002 ± 0.0004*** | -0.23 ± 0.06*** | -36.4 ± 14.4* |
| **Resting Y** | 0.72 ± 0.03*** | 0.31 ± 0.02*** | 0.42 ± 0.02*** | 0.58 ± 0.03*** | 0.26 ± 0.02*** | 0.55 ± 0.02*** | 0.96 ± 0.03*** |
| **BP Medication** | 0.55 ± 0.74 | 2.32 ± 0.43*** | 1.95 ± 0.40*** | 0.34 ± 0.79 | -0.009 ± 0.004** | 1.66 ± 0.58** | 330.0 ± 141.9* |
| **Smoking** | -0.13 ± 0.31 | 0.17 ± 0.18 | 0.08 ± 0.17 | -1.10 ± 0.35** | -0.002 ± 0.002 | -1.66 ± 0.25*** | -311.5 ± 61.2*** |
| **Alcohol** | 0.004 ± 0.01 | 0.02 ± 0.01* | 0.02 ± 0.01* | -0.002 ± 0.01 | -0.0001 ± 0.0001 | -0.0001 ± 0.01 | 2.32 ± 2.63 |
| **BMI** | 0.74 ± 0.06*** | 0.23 ± 0.03*** | 0.42 ± 0.03*** | 0.49 ± 0.06*** | 0.001 ± 0.0003*** | 1.12 ± 0.05*** | 230.2 ± 11.1*** |
| **METs×ACFE** | 0.04 ± 0.02* | -0.001 ± 0.01 | 0.02 ± 0.01 | 0.05 ± 0.02* | 0.0002 ± 0.0001* | 0.06 ± 0.01*** | 10.2 ± 4.5* |
| **METs×Sex** | -0.35 ± 0.08*** | 0.34 ± 0.05*** | 0.14 ± 0.05** | -0.61 ± 0.09*** | 0.0006 ± 0.0004<sup>#</sup> | 0.44 ± 0.06*** | -126.6 ± 17.5*** |
| **METs×Race** | -0.86 ± 0.08*** | 0.21 ± 0.05*** | -0.14 ± 0.05** | -1.02 ± 1.0*** | -0.003 ± 0.0004*** | -0.20 ± 0.06*** | -168.3 ± 17.7*** |

Abbreviations: ACFE, adverse childhood family environment; BMI, body mass index; BP, blood pressure.

**Supplementary Table 2.** Full results for the two-way mixed effects restricted maximum likelihood models conducted in sensitivity analyses. β coefficients ± SE are shown for each model term. In addition to the covariates from the primary models, sensitivity models were adjusted for waist circumference, exercise test duration, maximal heart rate, and two-minute heart-rate recovery. Models were also adjusted for study center (not shown). Because METs were centered at the lowest exercise stage (4.1 METs), the ACFE main effect reflects the association between ACFE exposure and the hemodynamic outcome at the lowest workload, and the METs×ACFE interaction term reflects the degree to which the ACFE effect changes with each additional MET of exercise intensity. ^#^, p < 0.1; *, *p* < 0.05; ** *p* < 0.01; \*\*\**p* < 0.001.

|  | SBP | DBP | MBP | PP | PPI | HR | RPP |
| --- | --- | --- | --- | --- | --- | --- | --- |
| **Intercept** | 148.1 ± 0.8*** | 76.6 ± 0.46*** | 100.2 ± 0.43*** | 72.9 ± 0.86*** | 0.49 ± 0.004*** | 110.8 ± 0.44*** | 16349.0 ± 137.3*** |
| **METs** | 6.00 ± 0.07*** | -0.73 ± 0.05*** | 1.51 ± 0.04*** | 6.67 ± 0.09*** | 0.02 ± 0.0003*** | 8.49 ± 0.05*** | 2319.0 ± 16.2*** |
| **ACFE** | -0.27 ± 0.14<sup>#</sup> | -0.15 ± 0.08<sup>#</sup> | -0.22 ± 0.08** | -0.10 ± 0.15 | 0.0001 ± 0.0007 | -0.40 ± 0.10*** | -75.3 ± 24.6** |
| **SEX** | -7.16 ± 0.92*** | -3.91 ± 0.53*** | -5.64 ± 0.50*** | -4.78 ± 1.0*** | -0.01 ± 0.005** | 1.53 ± 0.48* | -291.9 ± 156.4* |
| **RACE** | 0.17 ± 0.61 | -1.08 ± 0.35** | -0.85 ± 0.34* | 0.34 ± 0.66 | 0.006 ± 0.003<sup>#</sup> | 0.37 ± 0.42 | -204.2 ± 107.1<sup>#</sup> |
| **Age** | -0.09 ± 0.07 | 0.20 ± 0.04*** | 0.11 ± 0.04** | -0.19 ± 0.08* | -0.002 ± 0.0004*** | -0.08 ± 0.03** | -19.0 ± 12.7<sup>#</sup> |
| **Resting Y** | 0.69 ± 0.03*** | 0.30 ± 0.02*** | 0.40 ± 0.02*** | 0.58 ± 0.03*** | 0.26 ± 0.02*** | 0.06 ± 0.007*** | 0.53 ± 0.03*** |
| **BP Medication** | 0.09 ± 0.74 | 2.12 ± 0.43*** | 1.68 ± 0.40*** | -0.04 ± 0.80 | -0.009 ± 0.004** | -0.02 ± 0.27 | 332.7 ± 125.0** |
| **Smoking** | 0.26 ± 0.32 | 0.14 ± 0.19 | 0.17 ± 0.18 | -0.51 ± 0.36 | -0.007 ± 0.002 | -0.67 ± 0.11*** | -161.0 ± 55.0** |
| **Alcohol** | -0.003 ± 0.01 | 0.02 ± 0.008* | 0.02 ± 0.008* | -0.008 ± 0.02 | -0.0001 ± 0.0001<sup>#</sup> | -0.01 ± 0.005* | 3.62 ± 2.35 |
| **BMI** | 0.27 ± 0.13* | 0.11 ± 0.08 | 0.17 ± 0.07* | 0.10 ± 0.14 | 0.0002 ± 0.0006 | 0.16 ± 0.05** | 97.7 ± 22.3*** |
| **Waist Circumference** | 0.17 ± 0.06** | 0.02 ± 0.04 | 0.07 ± 0.03* | 0.17 ± 0.07* | 0.0006 ± 0.0003<sup>#</sup> | 0.002 ± 0.02 | 22.3 ± 10.3<sup>#</sup> |
| **GXT Duration** | -0.01 ± 0.003*** | -0.008 ± 0.002*** | -0.01 ± 0.001*** | -0.005 ± 0.003<sup>#</sup> | 0.000003 ± 0.00001 | -0.10 ± 0.001*** | -12.5 ± 0.47*** |
| **Maximum Heart Rate** | 0.18 ± 0.02*** | 0.04 ± 0.01*** | 0.09 ± 0.01*** | 0.17 ± 0.02*** | 0.0004 ± 0.0001*** | 0.91 ± 0.008*** | 114.1 ± 3.7*** |
| **Heart Rate Recovery** | -0.02 ± 0.02* | 0.007 ± 0.01 | -0.002 ± 0.01 | -0.06 ± 0.03*** | -0.0002 ± 0.0001<sup>#</sup> | -0.14 ± 0.008*** | -33.56 ± 4.19*** |
| **METs×ACFE** | 0.04 ± 0.02* | -0.001 ± 0.01 | 0.02 ± 0.01 | 0.05 ± 0.03* | 0.0002 ± 0.0001* | 0.05 ± 0.01*** | 9.9 ± 4.5* |
| **METs×Sex** | -0.34 ± 0.08*** | 0.35 ± 0.05*** | 0.14 ± 0.05** | -0.61 ± 0.09*** | 0.0006 ± 0.0004<sup>#</sup> | 0.36 ± 0.05*** | -127.5 ± 17.5*** |
| **METs×Race** | -0.86 ± 0.08*** | 0.21 ± 0.05*** | -0.14 ± 0.05** | -1.02 ± 1.0*** | -0.003 ± 0.0004*** | -0.13 ± 0.05* | -167.4 ± 17.7*** |

Abbreviations: ACFE, adverse childhood family environment; BMI, body mass index; BP, blood pressure; GXT, graded exercise test.

**Supplementary Figure 1.**
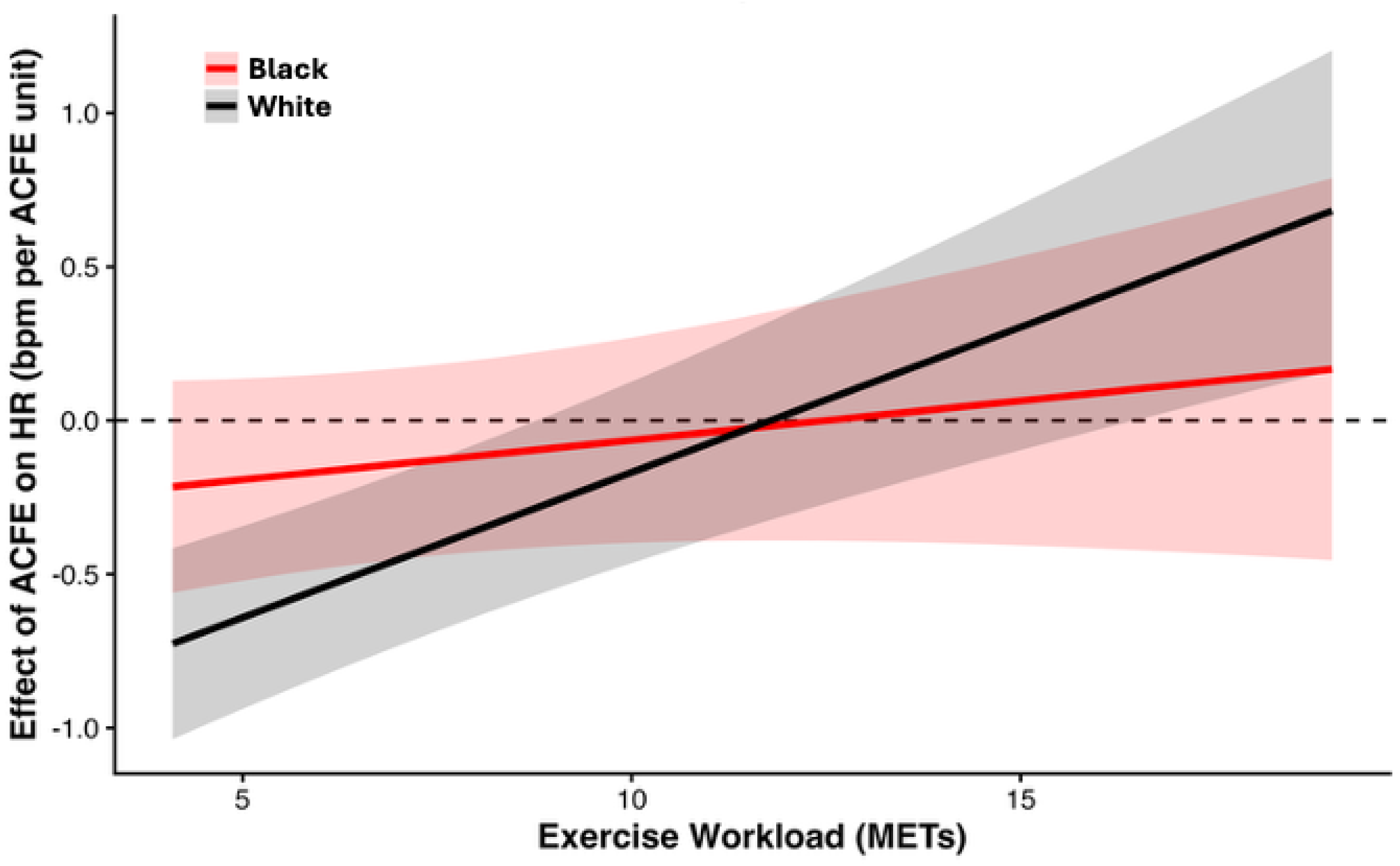
Race-stratified Marginal Effects of Adverse Childhood Family Environment (ACFE) on Heart Rate Responses during Graded Exercise. The figure shows the model-estimated marginal association of ACFE with heart rate (HR; beats·min⁻¹ per 1-unit increase in ACFE) across exercise workload (METs) for White (red line) and Black (black line) participants. Shaded bands represent the 95% confidence intervals calculated using the delta method from the fixed-effects variance–covariance matrix. Positive values indicate larger increases in HR with increasing ACFE, whereas negative values indicate smaller HR responses associated with higher adversity exposure. Consistent with the significant METs × ACFE × race interaction (F_1,2988.8_ = 5.04; β = 0.069 ± 0.03; *p* = 0.025), the ACFE–HR relation became progressively more positive with increasing workload, with a steeper workload-dependent change observed among White participants than Black participants.

## BIBLIOGRAPHY

1. Nelson CA, Scott RD, Bhutta ZA, Harris NB, Danese A, Samara M. Adversity in childhood is linked to mental and physical health throughout life. BMJ. 2020;371:m3048. doi: 10.1136/bmj.m3048

2. Bakema MJ, van Zuiden M, Collard D, Zantvoord JB, de Rooij SR, Elsenburg LK, Snijder MB, Stronks K, van den Born BH, Lok A. Associations Between Child Maltreatment, Autonomic Regulation, and Adverse Cardiovascular Outcome in an Urban Population: The HELIUS Study. Front Psychiatry. 2020;11:69. doi: 10.3389/fpsyt.2020.00069

3. Pierce JB, Kershaw KN, Kiefe CI, Jacobs DR, Jr., Sidney S, Merkin SS, Feinglass J. Association of Childhood Psychosocial Environment With 30-Year Cardiovascular Disease Incidence and Mortality in Middle Age. J Am Heart Assoc. 2020;9:e015326. doi: 10.1161/JAHA.119.015326

4. Danese A, Moffitt TE, Harrington H, Milne BJ, Polanczyk G, Pariante CM, Poulton R, Caspi A. Adverse childhood experiences and adult risk factors for age-related disease: depression, inflammation, and clustering of metabolic risk markers. Arch Pediatr Adolesc Med. 2009;163:1135–1143. doi: 10.1001/archpediatrics.2009.214

5. Dong M, Giles WH, Felitti VJ, Dube SR, Williams JE, Chapman DP, Anda RF. Insights into causal pathways for ischemic heart disease: adverse childhood experiences study. Circulation. 2004;110:1761–1766. doi: 10.1161/01.CIR.0000143074.54995.7F

6. Gilbert LK, Breiding MJ, Merrick MT, Thompson WW, Ford DC, Dhingra SS, Parks SE. Childhood adversity and adult chronic disease: an update from ten states and the District of Columbia, 2010. Am J Prev Med. 2015;48:345-349. doi: 10.1016/j.amepre.2014.09.006

7. Jenkins NDM, Rogers EM, Banks NF, Tomko PM, Sciarrillo CM, Emerson SR, Taylor A, Teague TK. Childhood psychosocial stress is linked with impaired vascular endothelial function, lower SIRT1, and oxidative stress in young adulthood. Am J Physiol Heart Circ Physiol. 2021;321:H532–H541. doi: 10.1152/ajpheart.00123.2021

8. Rodriguez-Miguelez P, Looney J, Blackburn M, Thomas J, Pollock JS, Harris RA. The Link Between Childhood Adversity and Cardiovascular Disease Risk: Role of Cerebral and Systemic Vasculature. Function (Oxf). 2022;3:zqac029. doi: 10.1093/function/zqac029

9. Jenkins NDM, Robinson AT. How do Adverse Childhood Experiences get Under the Skin to Promote Cardiovascular Disease? A Focus on Vascular Health. Function. 2022;3:zqac032. doi: 10.1093/function/zqac032

10. Su S, Wang X, Pollock JS, Treiber FA, Xu X, Snieder H, McCall WV, Stefanek M, Harshfield GA. Adverse childhood experiences and blood pressure trajectories from childhood to young adulthood: the Georgia stress and Heart study. Circulation. 2015;131:1674–1681. doi: 10.1161/CIRCULATIONAHA.114.013104

11. Tzemos N, Lim PO, MacDonald TM. Is exercise blood pressure a marker of vascular endothelial function? QJM. 2002;95:423–429. doi: 10.1093/qjmed/95.7.423

12. Allison TG, Cordeiro MA, Miller TD, Daida H, Squires RW, Gau GT. Prognostic significance of exercise-induced systemic hypertension in healthy subjects. Am J Cardiol. 1999;83:371–375. doi: 10.1016/s0002-9149(98)00871-6

13. Filipovsky J, Ducimetiere P, Safar ME. Prognostic significance of exercise blood pressure and heart rate in middle-aged men. Hypertension. 1992;20:333–339. doi: 10.1161/01.hyp.20.3.333

14. Mundal R, Kjeldsen SE, Sandvik L, Erikssen G, Thaulow E, Erikssen J. Exercise blood pressure predicts cardiovascular mortality in middle-aged men. Hypertension. 1994;24:56–62. doi: 10.1161/01.hyp.24.1.56

15. Weiss SA, Blumenthal RS, Sharrett AR, Redberg RF, Mora S. Exercise blood pressure and future cardiovascular death in asymptomatic individuals. Circulation. 2010;121:2109–2116. doi: 10.1161/CIRCULATIONAHA.109.895292

16. Zanstra YJ, Johnston DW. Cardiovascular reactivity in real life settings: measurement, mechanisms and meaning. Biol Psychol. 2011;86:98–105. doi: 10.1016/j.biopsycho.2010.05.002

17. Taylor TR, Kamarck TW, Dianzumba S. Cardiovascular reactivity and left ventricular mass: an integrative review. Ann Behav Med. 2003;26:182–193. doi: 10.1207/S15324796ABM2603_03

18. Bourassa KJ, Moffitt TE, Harrington H, Houts R, Poulton R, Ramrakha S, Caspi A. Lower Cardiovascular Reactivity is Associated with More Childhood Adversity and Poorer Midlife Health: Replicated Findings from the Dunedin and MIDUS Cohorts. Clin Psychol Sci. 2021;9:961–978. doi: 10.1177/2167702621993900

19. Rogers EM, Banks NF, Tomko PM, Sciarrillo CM, Emerson SR, Thomas EBK, Taylor A, Teague TK, Jenkins NDM. Progressive exercise training improves cardiovascular psychophysiological outcomes in young adult women with a history of adverse childhood experiences. J Appl Physiol (1985). 2023;134:742–752. doi: 10.1152/japplphysiol.00524.2022

20. Gobel FL, Norstrom LA, Nelson RR, Jorgensen CR, Wang Y. The rate-pressure product as an index of myocardial oxygen consumption during exercise in patients with angina pectoris. Circulation. 1978;57:549–556. doi: 10.1161/01.cir.57.3.549

21. Boutouyrie P, Chowienczyk P, Humphrey JD, Mitchell GF. Arterial Stiffness and Cardiovascular Risk in Hypertension. Circ Res. 2021;128:864–886. doi: 10.1161/CIRCRESAHA.121.318061

22. Franklin SS. Ageing and hypertension: the assessment of blood pressure indices in predicting coronary heart disease. J Hypertens Suppl. 1999;17:S29–36.

23. Soares AL, Howe LD, Matijasevich A, Wehrmeister FC, Menezes AM, Goncalves H. Adverse childhood experiences: Prevalence and related factors in adolescents of a Brazilian birth cohort. Child Abuse Negl. 2016;51:21–30. doi: 10.1016/j.chiabu.2015.11.017

24. Carpenter T, Grecian SM, Reynolds RM. Sex differences in early-life programming of the hypothalamic-pituitary-adrenal axis in humans suggest increased vulnerability in females: a systematic review. J Dev Orig Health Dis. 2017;8:244–255. doi: 10.1017/S204017441600074X

25. Suglia SF, Koenen KC, Boynton-Jarrett R, Chan PS, Clark CJ, Danese A, Faith MS, Goldstein BI, Hayman LL, Isasi CR, et al. Childhood and Adolescent Adversity and Cardiometabolic Outcomes: A Scientific Statement From the American Heart Association. Circulation. 2018;137:e15–e28. doi: 10.1161/CIR.0000000000000536

26. Lehman BJ, Taylor SE, Kiefe CI, Seeman TE. Relation of childhood socioeconomic status and family environment to adult metabolic functioning in the CARDIA study. Psychosom Med. 2005;67:846–854. doi: 10.1097/01.psy.0000188443.48405.eb

27. Felitti VJ, Anda RF, Nordenberg D, Williamson DF, Spitz AM, Edwards V, Koss MP, Marks JS. Relationship of Childhood Abuse and Household Dysfunction to Many of the Leading Causes of Death in Adults: The Adverse Childhood Experiences (ACE) Study. American Journal of Preventive Medicine. 1998;14:245–258. doi: 10.1016/S0749-3797(98)00017-8

28. Pierce J, Kershaw K, Kiefe C, Jacobs D, Jr., Sidney S, Merkin S, Feinglass J. Association of Childhood Psychosocial Environment With 30-Year Cardiovascular Disease Incidence and Mortality in Middle Age. J Am Heart Assoc. 2020;9:e015326. doi: 10.1161/JAHA.119.015326

29. Merrick MT, Ford DC, Ports KA, Guinn AS. Prevalence of Adverse Childhood Experiences From the 2011-2014 Behavioral Risk Factor Surveillance System in 23 States. JAMA Pediatr. 2018;172:1038–1044. doi: 10.1001/jamapediatrics.2018.2537

30. Swedo EA, Aslam MV, Dahlberg LL, Niolon PH, Guinn AS, Simon TR, Mercy JA. Prevalence of Adverse Childhood Experiences Among U.S. Adults - Behavioral Risk Factor Surveillance System, 2011-2020. MMWR Morb Mortal Wkly Rep. 2023;72:707-715. doi: 10.15585/mmwr.mm7226a2

31. Miyai N, Arita M, Miyashita K, Morioka I, Shiraishi T, Nishio I. Blood pressure response to heart rate during exercise test and risk of future hypertension. Hypertension. 2002;39:761–766. doi: 10.1161/hy0302.105777

32. Singh JP, Larson MG, Manolio TA, O’Donnell CJ, Lauer M, Evans JC, Levy D. Blood pressure response during treadmill testing as a risk factor for new-onset hypertension. The Framingham heart study. Circulation. 1999;99:1831–1836. doi: 10.1161/01.cir.99.14.1831

33. Dlin RA, Hanne N, Silverberg DS, Bar-Or O. Follow-up of normotensive men with exaggerated blood pressure response to exercise. Am Heart J. 1983;106:316–320. doi: 10.1016/0002-8703(83)90198-9

34. Manolio TA, Burke GL, Savage PJ, Sidney S, Gardin JM, Oberman A. Exercise blood pressure response and 5-year risk of elevated blood pressure in a cohort of young adults: the CARDIA study. Am J Hypertens. 1994;7:234–241. doi: 10.1093/ajh/7.3.234

35. Matthews CE, Pate RR, Jackson KL, Ward DS, Macera CA, Kohl HW, Blair SN. Exaggerated blood pressure response to dynamic exercise and risk of future hypertension. J Clin Epidemiol. 1998;51:29–35. doi: 10.1016/s0895-4356(97)00223-0

36. Lane-Cordova AD, Carnethon MR, Catov JM, Montag S, Lewis CE, Schreiner PJ, Dude A, Sternfeld B, Badon SE, Greenland P, et al. Cardiorespiratory fitness, exercise haemodynamics and birth outcomes: the Coronary Artery Risk Development in Young Adults Study. BJOG. 2018;125:1127–1134. doi: 10.1111/1471-0528.15146

37. McLaughlin KA, Sheridan MA, Alves S, Mendes WB. Child maltreatment and autonomic nervous system reactivity: identifying dysregulated stress reactivity patterns by using the biopsychosocial model of challenge and threat. Psychosom Med. 2014;76:538–546. doi: 10.1097/PSY.0000000000000098

38. Winzeler K, Voellmin A, Hug E, Kirmse U, Helmig S, Princip M, Cajochen C, Bader K, Wilhelm FH. Adverse childhood experiences and autonomic regulation in response to acute stress: the role of the sympathetic and parasympathetic nervous systems. Anxiety Stress Coping. 2017;30:145–154. doi: 10.1080/10615806.2016.1238076

39. Gooding HC, Milliren CE, Austin SB, Sheridan MA, McLaughlin KA. Child Abuse, Resting Blood Pressure, and Blood Pressure Reactivity to Psychosocial Stress. J Pediatr Psychol. 2016;41:5–14. doi: 10.1093/jpepsy/jsv040

40. Dempster KS, Wade TJ, MacNeil AJ, O’Leary DD. Adverse childhood experiences are associated with altered cardiovascular reactivity to head-up tilt in young adults. Am J Physiol Regul Integr Comp Physiol. 2023;324:R425–R434. doi: 10.1152/ajpregu.00148.2022

41. Zhong X, Ming Q, Dong D, Sun X, Cheng C, Xiong G, Li C, Zhang X, Yao S. Childhood Maltreatment Experience Influences Neural Response to Psychosocial Stress in Adults: An fMRI Study. Front Psychol. 2019;10:2961. doi: 10.3389/fpsyg.2019.02961

42. Heim C, Newport DJ, Heit S, Graham YP, Wilcox M, Bonsall R, Miller AH, Nemeroff CB. Pituitary-adrenal and autonomic responses to stress in women after sexual and physical abuse in childhood. JAMA. 2000;284:592–597. doi: 10.1001/jama.284.5.592

43. Treiber FA, Kamarck T, Schneiderman N, Sheffield D, Kapuku G, Taylor T. Cardiovascular reactivity and development of preclinical and clinical disease states. Psychosom Med. 2003;65:46–62. doi: 10.1097/00006842-200301000-00007

44. Smith TW, Nealey JB, Kircher JC, Limon JP. Social determinants of cardiovascular reactivity: effects of incentive to exert influence and evaluative threat. Psychophysiology. 1997;34:65–73. doi: 10.1111/j.1469-8986.1997.tb02417.x

45. Cohen S, Janicki-Deverts D, Chen E, Matthews KA. Childhood socioeconomic status and adult health. Ann N Y Acad Sci. 2010;1186:37–55. doi: 10.1111/j.1749-6632.2009.05334.x

46. Matthews KA, Salomon K, Brady SS, Allen MT. Cardiovascular reactivity to stress predicts future blood pressure in adolescence. Psychosom Med. 2003;65:410–415. doi: 10.1097/01.psy.0000057612.94797.5f

47. Stewart KJ, Sung J, Silber HA, Fleg JL, Kelemen MD, Turner KL, Bacher AC, Dobrosielski DA, DeRegis JR, Shapiro EP, et al. Exaggerated exercise blood pressure is related to impaired endothelial vasodilator function. Am J Hypertens. 2004;17:314–320. doi: 10.1016/S0895-7061(03)01003-3

48. McEniery CM, Wallace S, Mackenzie IS, McDonnell B, Yasmin, Newby DE, Cockcroft JR, Wilkinson IB. Endothelial function is associated with pulse pressure, pulse wave velocity, and augmentation index in healthy humans. Hypertension. 2006;48:602–608. doi: 10.1161/01.HYP.0000239206.64270.5f

49. Rodriguez-Miguelez P, Looney J, Blackburn M, Thomas J, Pollock JS, Harris RA. The Link Between Childhood Adversity and Cardiovascular Disease Risk: Role of Cerebral and Systemic Vasculature. Function. 2022. doi: 10.1093/function/zqac029

50. Jenkins NDM, Robinson AT. How do Adverse Childhood Experiences get Under the Skin to Promote Cardiovascular Disease? A Focus on Vascular Health. Function (Oxf). 2022;3:zqac032. doi: 10.1093/function/zqac032

51. von Kanel R, Princip M, Holzgang SA, Giannopoulos AA, Kaufmann PA, Buechel RR, Zuccarella-Hackl C, Pazhenkottil AP. Cross-sectional study on the impact of adverse childhood experiences on coronary flow reserve in male physicians with and without occupational burnout. J Psychosom Res. 2024;181:111672. doi: 10.1016/j.jpsychores.2024.111672

52. Monge Garcia MI, Santos A. Understanding ventriculo-arterial coupling. Ann Transl Med. 2020;8:795. doi: 10.21037/atm.2020.04.10

53. Baral R, Loudon B, Frenneaux MP, Vassiliou VS. Ventricular-vascular coupling in heart failure with preserved ejection fraction: A systematic review and meta-analysis. Heart Lung. 2021;50:121–128. doi: 10.1016/j.hrtlng.2020.07.002

54. Chantler PD, Lakatta EG, Najjar SS. Arterial-ventricular coupling: mechanistic insights into cardiovascular performance at rest and during exercise. J Appl Physiol (1985). 2008;105:1342–1351. doi: 10.1152/japplphysiol.90600.2008

55. Brindle RC, Pearson A, Ginty AT. Adverse childhood experiences (ACEs) relate to blunted cardiovascular and cortisol reactivity to acute laboratory stress: A systematic review and meta-analysis. Neurosci Biobehav Rev. 2022;134:104530. doi: 10.1016/j.neubiorev.2022.104530

56. Ouellet-Morin I, Robitaille MP, Langevin S, Cantave C, Brendgen M, Lupien SJ. Enduring effect of childhood maltreatment on cortisol and heart rate responses to stress: The moderating role of severity of experiences. Dev Psychopathol. 2019;31:497–508. doi: 10.1017/S0954579418000123

57. Bobba-Alves N, Sturm G, Lin J, Ware SA, Karan KR, Monzel AS, Bris C, Procaccio V, Lenaers G, Higgins-Chen A, et al. Cellular allostatic load is linked to increased energy expenditure and accelerated biological aging. Psychoneuroendocrinology. 2023;155:106322. doi: 10.1016/j.psyneuen.2023.106322

58. Reid DM, Choe JY, Bruce MA, Thorpe RJ, Jones HP, Phillips NR. Mitochondrial Functioning: Front and Center in Defining Psychosomatic Mechanisms of Allostasis in Health and Disease. In: Yan Q, ed. Psychoneuroimmunology: Methods and Protocols. New York, NY: Springer US; 2025:91-110.

59. Bobba-Alves N, Juster RP, Picard M. The energetic cost of allostasis and allostatic load. Psychoneuroendocrinology. 2022;146:105951. doi: 10.1016/j.psyneuen.2022.105951

60. Picard M, Murugan NJ. The energy resistance principle. Cell Metab. 2025;37:2107–2127. doi: 10.1016/j.cmet.2025.09.002

